# *ACE2* gene variants may underlie interindividual variability and susceptibility to COVID-19 in the Italian population

**DOI:** 10.1101/2020.04.03.20047977

**Authors:** Benetti Elisa, Tita Rossella, Spiga Ottavia, Ciolfi Andrea, Birolo Giovanni, Bruselles Alessandro, Doddato Gabriella, Giliberti Annarita, Marconi Caterina, Musacchia Francesco, Pippucci Tommaso, Torella Annalaura, Trezza Alfonso, Valentino Floriana, Baldassarri Margherita, Brusco Alfredo, Asselta Rosanna, Bruttini Mirella, Furini Simone, Seri Marco, Nigro Vincenzo, Matullo Giuseppe, Tartaglia Marco, Mari Francesca, Renieri Alessandra, Pinto Anna Maria

## Abstract

In December 2019, an initial cluster of interstitial bilateral pneumonia emerged in Wuhan, China. A human-to-human transmission was assumed and a previously unrecognized entity, termed coronavirus-disease-19 (COVID-19) due to a novel coronavirus (SARS-CoV-2) was described. The infection has rapidly spread out all over the world and Italy has been the first European country experiencing the endemic wave with unexpected clinical severity in comparison with Asian countries.

It has been shown that SARS-CoV-2 utilizes angiotensin converting enzyme 2 (ACE2) as host receptor and host proteases for cell surface binding and internalization. Thus, a predisposing genetic background can give reason for inter-individual disease susceptibility and/or severity. Taking advantage of the Network of Italian Genomes (NIG), here we mined whole-exome-sequencing data of 6930 Italian control individuals from five different centers looking for *ACE2* variants. A number of variants with a potential impact on protein stability were identified. Among these, three more common missense changes, p.(Asn720Asp), p.(Lys26Arg), p.(Gly211Arg) were predicted to interfere with protein structure and stabilization. Rare variants likely interfering with the internalization process, namely p.(Leu351Val) and p.(Pro389His), predicted to interfere with SARS-CoV-2 spike protein binding, were also observed. Comparison of *ACE2* WES data between a cohort of 131 patients and 258 controls allowed identifying a statistically significant (P value <0,029) higher allelic variability in controls compared to patients. These findings suggest that a predisposing genetic background may contribute to the observed inter-individual clinical variability associated with COVID-19, allowing an evidence-based risk assessment leading to personalized preventive measures and therapeutic options.

## INTRODUCTION

In December 2019, a new infectious respiratory disease emerged in Wuhan, Hubei province, China (1–3). An initial cluster of infections likely due to animal-to-human transmission was rapidly followed by a human-to-human transmission (4). The disease was recognized to be caused by a novel coronavirus (SARS-CoV-2) and termed coronavirus disease 19 (COVID-19). The infection spread within China and all over the world, and it has been declared as pandemic by the World Health Organization (WHO) on 2^nd^ March 2020. The symptoms of COVID-19 range from fever, dry cough, fatigue, congestion, sore throat and diarrhea to severe interstitial bilateral pneumonia with a ground–glass image at the CT scan. While recent studies provide evidence of a high number of asymptomatic or paucisymptomatic patients who represent the main reservoir for the infection progression, the severe cases can rapidly evolve towards a respiratory distress syndrome which can be lethal (5). Although age and comorbidity have been described as the main determinants of disease progression towards severe respiratory distress, the high variation in clinical severity among middle-age adults and children would likely suggest a strong role of the host genetic asset.

A high sequence homology has been shown between SARS-associated coronavirus (SARS-CoV) and SARS-CoV-2 (6). Recent studies modelled the spike protein to identify the receptor for SARS-CoV-2 and indicated that angiotensin converting enzyme 2 (ACE2) is the receptor for this novel coronavirus(7,8). Zhou et. al. conducted virus infectivity studies and showed that ACE2 is essential for SARS-CoV-2 to enter HeLa cells (9). Although the binding strength between SARS-CoV-2 and ACE2 is weaker than that between SARS-CoV and ACE2, it is considered as much high as threshold necessary for virus infection. The spike glycoprotein (S-protein), a trimeric glycoprotein in the virion surface (giving the name of crown *-corona* in latin-), mediates receptor recognition throughout its receptor binding domain (RBD) and membrane fusion (10,11). Based on recent reports, SARS-CoV-2 protein binds to ACE2 through Leu455, Phe486, Gln493, Ala501 and Tyr505. It has been postulated that residues 31, 41, 82, 353, 355, and 357 of the ACE2 receptor map to the surface of the protein interacting with SARS-CoV-2 spike protein (12), as previously documented for SARS-CoV. Following interaction, cleavage of the C-terminal segment of ACE2 by proteases, such as transmembrane protease serine 2 (TMPRSS2), enhances the spike protein-driven viral entry (13,14). Thus, it is possible, in principle, that genetic variability of the ACE2 receptor is one of the elements modulating virion intake and thus disease severity. *ACE2* is located on chromosome X. Although it is one of the genes escaping X inactivation several lines of evidence suggest that a different degree of X-chromosome inactivation is present in distinct tissues (15).

Taking advantage of the Network of Italian Genomes (NIG), a consortium established to generate a public database (NIG-db) containing aggregate variant frequencies data for the Italian population (http://www.nig.cineca.it/), here we describe the genetic variation of *ACE2* in the Italian population, one of the newly affected countries by the SARS-CoV-2 outbreak causing COVID-19. Three common c.2158A>G p.(Asn720Asp), c.77A>G p.(Lys26Arg) and c.631G>A p.(Gly211Arg) variants and 27 rare missense variants were identified, 9 of which had not previously been reported in public databases. We show that p.(Asn720Asp), which lies in a residue located close to the cleavage sequence of TMPRSS2, likely affects the cleavage-dependent virion intake. Along with the other two common variants, this substitution is represented in the Italian and European populations but is extremely rare in the Asian population. We also show that two rare variants, namely, c.1051C>G p.(Leu351Val) and c.1166C>A p.(Pro389His) are predicted to cause conformational changes impacting RBD interaction. As the uncertainty regarding the transmissibility and severity of disease rise, we believe that a deeper characterisation of the host genetics and functional characterization of variants may help not only in understanding the pathophysiology of the disease but also in envisaging risk assessment.

### Materials and Methods

#### Italian Population randomization

The work has been realized in the context of the Network of Italian Genomes (NIG), with the contribution of centers: Azienda Ospedaliera Universitaria Senese, Azienda Ospedaliera-Universitaria Policlinico Sant’Orsola-Malpighi di Bologna, Città della Salute e della Scienza di Torino, Università della Campania “Luigi Vanvitelli”, Ospedale Pediatrico Bambino Gesù. The NIG (http://www.nig.cineca.it/) aim is to create a shared database (NIG-db) containing data from nucleic acids sequencing of Italian subjects. This database allows defining an Italian Reference Genome for the identification of genes responsible for genetic diseases or Italian population susceptibility to complex disorders and for the detection of genetic variants responsible for interindividual differences in disease progression ad /or drug response among the Italian population. Individuals coming to our centers were offered to participate to the NIG study and blood withdrawal was performed upon informed consent. Individuals provided signed informed consents at each participating center for whole exome sequencing analysis (WES), and clinical and molecular data storage and usage. All subjects were unrelated, healthy, and of Italian ancestry. Italian origin was ascertained asking for parents and grandparents origin. DNA has been stored in the Telethon Network of Genetic Biobanks (project no. GTB12001), funded by Telethon Italy.

#### COVID-19 patients enrollment

The study was consistent with Institutional guidelines and approved by the University Hospital (Azienda Ospedaliera Universitaria Senese) Ethical Committee, Siena, Italy (Prot n. 16929, dated March 16, 2020). Written informed consent was obtained from all patients and controls. Peripheral blood samples in EDTA-containing tubes and detailed clinical data were collected. All these data were inserted in a section of the established and certified Biobank and Registry of the Medical Genetics Unit of the Hospital dedicated to COVID-19. The cohort of COVID-19 patients consists of 131 individuals out of whom 34 females and 97 males belonging to the GEN-COVID MULTICENTER STUDY (16, Late Breaking Abstract ESHG 2020.2 Virtual Conference “WES profiling of COVID-19”). The cohort of controls consists of 258 italian individuals (129 males and 129 females). All patients are of Italian ethnicity. The median age is 64 years (range 31-98): median age for women 66 years and for males 63 years. The population was clustered into four qualitative severity groups depending on the respiratory impairment and the need for ventilation: high care intensity group (those requiring invasive ventilation), intermediate care intensity group (those requiring non-invasive ventilation i.e. CPAP and BiPAP, and high-flows oxygen therapy), low care intensity group (those requiring conventional oxygen therapy) and very low care intensity group (those not requiring oxygen therapy).

#### Whole Exome Sequencing

Targeted enrichment and massively parallel sequencing were performed on genomic DNA extracted from circulating leukocytes of 6930 individuals. Genomic DNA was extracted from peripheral blood samples using standard procedures. Exome capture was carried out using SureSelect Human All Exon V4/V5/V6/V7 (Agilent Technologies, Santa Clara, CA), Clinical Research Exome V1/V2 (Agilent), Nextera Rapid Capture v.1.2 (Illumina, San Diego, CA), TruSeq Exome Targeted Regions (Illumina, San Diego, CA), TruSight One Expanded V2 (Illumina, San Diego, CA), Sequencing-by-Synthesis Kit v3/v4 (Illumina, San Diego, CA) or HiSeq 2000 v2 Sequencing-by-Synthesis Kit (Illumina, San Diego, CA) and sequencing was performed on Genome Analyzer (v3/v4)/HiSeq2000/NextSeq550/NextSeq500/Novaseq6000 platforms (Illumina, San Diego, CA). A subset of WES had been outsourced (BGI, Shenzen China; Mount Sinai, New York, USA; Broad Institute, Harvard, USA). Alignment of raw reads against reference genome Hg19,variant calling and annotation were attained using in-house pipelines (17-19) which take advantage of the GATK Best Practices workflow (20) and of Annovar, VEP (21,22). The genome aggregation database gnomAD (https://gnomad.broadinstitute.org/) was used to assess allele frequency for each variant among different populations. The mean depth of coverage of each *ACE2* exon in all participants was 55X. Variants with a depth of coverage lower that 20X were filtered out according to ASHG Guidelines for germline variants. (23).

The identified variants have been submitted in LOVD database:

Variant ID 0000667129 https://databases.lovd.nl/shared/individuals/00302622;

Variant ID 0000667137 https://databases.lovd.nl/shared/individuals/00302630;

Variant ID 0000667136 https://databases.lovd.nl/shared/individuals/00302628

Variant ID 0000667138 https://databases.lovd.nl/shared/individuals/00302629;

Variant ID 0000667131 https://databases.lovd.nl/shared/individuals/00302624;

Variant ID 0000667133 https://databases.lovd.nl/shared/individuals/00302626;

Variant ID 0000667130 https://databases.lovd.nl/shared/individuals/00302621;

Variant ID 0000667134 https://databases.lovd.nl/shared/individuals/00302625;

Variant ID 0000667132 https://databases.lovd.nl/shared/individuals/00302623;

Variant ID 0000667128 https://databases.lovd.nl/shared/individuals/00302620;

Variant ID 0000667126 https://databases.lovd.nl/shared/individuals/00302618;

Variant ID 0000667127 https://databases.lovd.nl/shared/individuals/00302619;

Variant ID 0000667125 https://databases.lovd.nl/shared/individuals/00302617;

Variant ID 0000667123 https://databases.lovd.nl/shared/individuals/00302615;

Variant ID 0000667124 https://databases.lovd.nl/shared/individuals/00302616;

Variant ID 0000667118 https://databases.lovd.nl/shared/individuals/00302610;

Variant ID 0000667120 https://databases.lovd.nl/shared/individuals/00302612;

Variant ID 0000667122 https://databases.lovd.nl/shared/individuals/00302614;

Variant ID 0000667121 https://databases.lovd.nl/shared/individuals/00302613;

Variant ID 0000667119 https://databases.lovd.nl/shared/individuals/00302611;

Variant ID 0000667117 https://databases.lovd.nl/shared/individuals/00302609;

#### Computational studies

The structure of native human angiotensin converting enzyme-related carboxypeptidase (ACE2) was downloaded from Protein Data Bank (https://www.rcsb.org/) (PDB ID code 1R42) (24). The DUET program (25) was used to predict the possible effect of amino acids substitutions on the protein structure and function, based on the use of machine-learning algorithms exploiting the three dimensional structure to quantitatively predict the effects of residue substitutions on protein functionality. Molecular dynamics (MD) simulations of wild-type and variant ACE2 proteins were carried out in GROMACS 2019.3 (26) to calculate root-mean-square deviation (RMSD) to define structural stability. The graphs were plotted by the XMGrace software (27). MD simulations were performed using a high parallel computing infrastructure (HPCS) with 660 cpu within 21 different nodes, 190T of RAM, 30T hard disk partition size and 6 NVIDIA TESLA gpu with CUDA support. PyMOL2.3 was used as a molecular graphic interface. The protein structures were solvated in a triclinic box filled with TIP3P water molecules and Na +/Cl - ions were added to neutralize the system. The whole systems were then minimized with a maximal force tolerance of 1000 kJ mol-1 nm-1 using the steepest descendent algorithm. The optimized systems were gradually heated to 310 K in 1 ns in the NVT ensemble, followed by 10 ns equilibration in the NPT ensemble at 1 atm and 310 K, using the V-Rescale thermostat and Berendsen barostat (28,29). Subsequently, a further 100 ns MD simulations were performed for data analysis.

## RESULTS

### ACE2 variants identification

The extent of variability along the entire *ACE2* coding sequence and flanking intronic stretches was assessed using 6930 Italian WES, out of which 4171 males and 2759 females which sum up to 9689 alleles. Identified variants and predicted effects on protein stability are summarized in Table 1, Table S1 and Table 2, and represented in Fig.1. Three more common variants, c.2158A>G p.(Asn720Asp), c.77A>G p.(Lys26Arg), c.631G>A p.(Gly211Arg), were identified. The c.2158A>G p.(Asn720Asp) substitution was estimated to have a frequency of 0.011 (103/9689 alleles), which is in line with the frequency of the variant reported in the gnomAD database (0.016), and is lower than the frequency reported in gnomAD for the European non-Finnish population (0.025, 2195/87966 analyzed alleles). Given the *ACE2* localization on X chromosome we focused our attention on the females alleles. All analysed females (2759 out of 6930) belonging to the Italian population, were heterozygotes for the variant. Notably, this variant has not been reported in the Eastern Asia population (13,784 exomes). The c.77A>G p.(Lys26Arg), c.631G>A p.(Gly211Arg) variants were found with a frequency of 0.0011 (lower than the frequency in the European-non Finnish population, 0.0058) and 0.0012 (European-non Finnish population frequency, 0.0019), respectively. Out of ~92708 analyzed alleles in the European non-Finnish population, one homozygous female has been reported for the c.77A>G p.(Lys26Arg) while no homozygous females were reported for the c.631G>A p.(Gly211Arg). According to gnomAD, the allele frequency of the c.77A>G p.(Lys26Arg) variant in the Eastern Asia population was 6×10^-5^, while the c.631G>A p.(Gly211Arg) has not been reported in 14.822 exomes. In addition to these variants, 28 rare missense variants were identified, out of which 10 had not previously been reported in GnomAD database and 9 truncating variants that had not been reported in gnomAD database (Table 1, Supplementary Table 1). Out of these variants, two fall in the neck domain, which is essential for dimerization and one in the intracellular domain. Many of them truncate the protein in different positions of the Protease domain embedded in the extracellular domain, which contains the receptor binding site for SARS-CoV-2. Only three truncating variants have been previously described for *ACE2* likely due to a low-tolerance for loss-of function variants. In line with this evidence, all these variants were very rare and no homozygous females were detected for the identified variants. Three missense changes c.1517T>C p.(Val506Ala), c.626T>G p.(Val209Gly), c.1129G>T p.(Gly377Glu) were predicted to have destabilizing structural consequences (Table 2); among these, c.1517T>C p.(Val506Ala) is indeed the only amino acid change reported in the European non-Finnish population (rs775181355; allele frequency 1.40×10^-5^, CADD 27,2) and is predicted as probably damaging for the protein structure by Polyphen and deleterious by SIFT. Similarly, c.1051C>G p.(Leu351Val) and c.1166C>A p.(Pro389His), which affect a highly hydrophobic core, were predicted to induce conformational changes influencing the interaction with spike protein. The amino acid substitution c.1166C>A p.(Pro389His) (rs762890235, European-non Finnish population allele frequency: 2.45 10^-5, CADD 24,8) was predicted to be probably damaging by Polyphen and deleterious by SIFT. Moreover, this rare variant has never been reported in Asian populations.

**Table 1.**
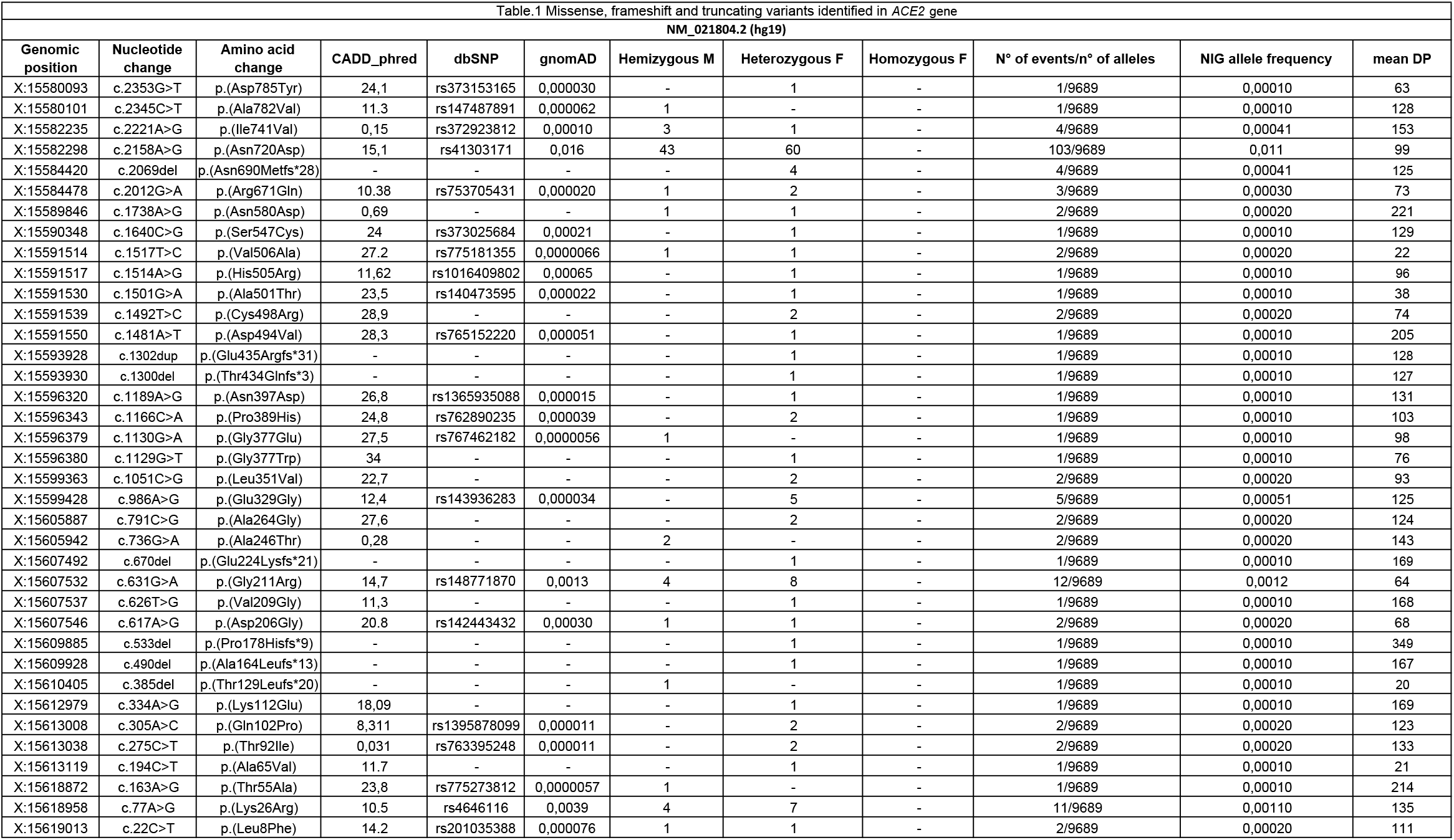
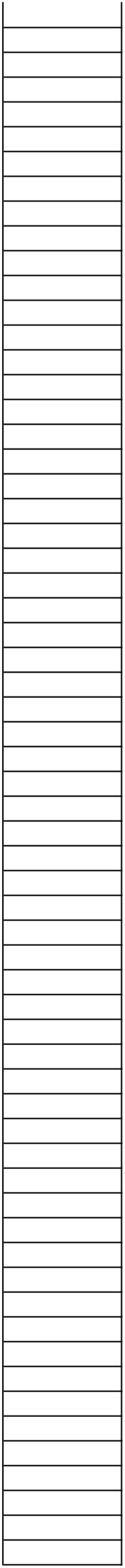
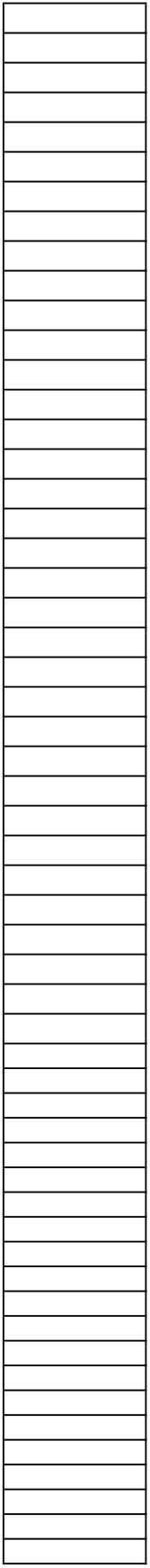
Missense, frameshift and truncating variants identified in *ACE2* gene. The table reports the genomic position, the nucleotidic and protein change of exonic *ACE2* identified variants. The genomic reference sequence is NM_021804.2 (hg19). CADD_phred scores are reported for the missense variants. When available, dbSNP rs number and the genome aggregation database gnomAD allele frequency are reported. For all variants are reported the number of individuals hemizygous, heterozygous or homozygous.

**Table 2.**
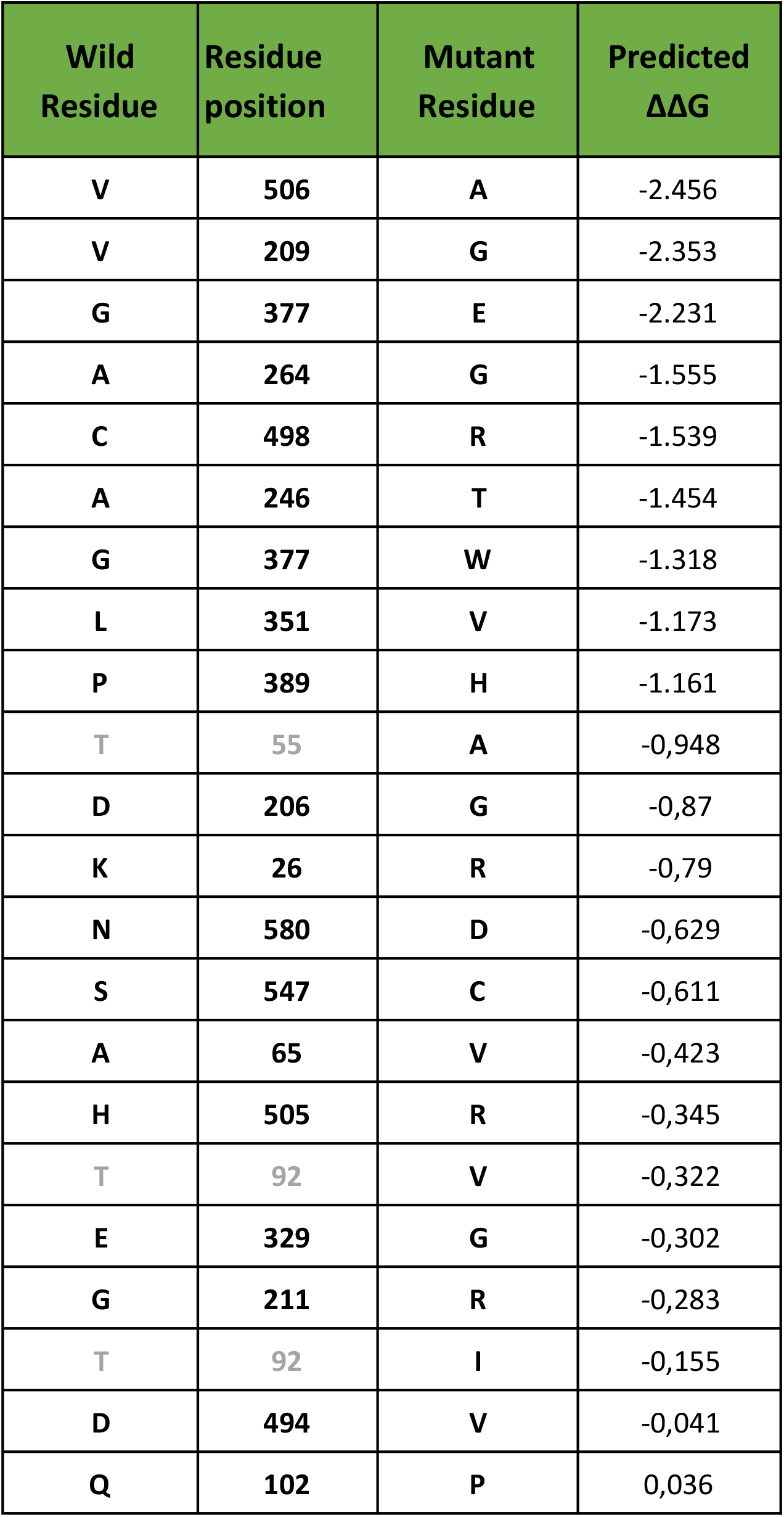
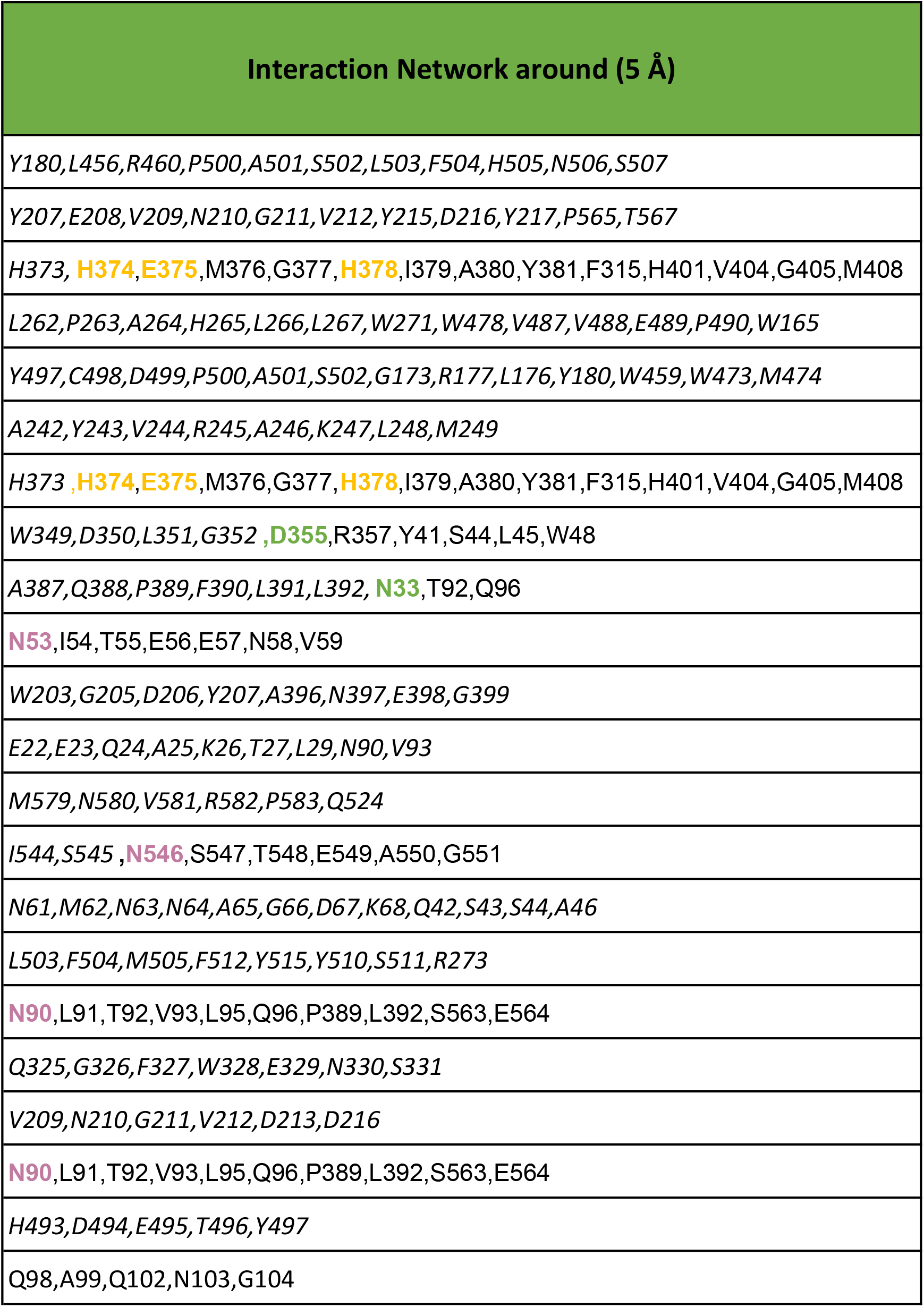
Predicted changes in ACE2 protein stability as consequence of residues changes. DUET program results that display predicted change in folding free energy upon ACE2 missense variant (∆∆G in kcal/mol). In the first three columns are reported single missense variants with specific position on ACE2 protein. The residues in the first column highlighted in grey are involved in N-glycosylation pattern NxT/S, therefore those missense variants determine the loss glycosylation of Asparagine 53 and 90 respectively. In the fourth column is reported G analysis predict effects of missense variants on protein stability using an integrated computational approach. The column “Interaction Network around (5 Å)” shows for each single missense variant the residues around 5Å. In this column, we highlight in green residues involved in spike SARS-CoV protein interaction, in yellow residues involved in Zinc coordination and finally in magenta residues of Asn involved in N-glycosylation. The last column defines the outcome of protein stability for each single missense variant. An increasing negative value for the ∆∆G is correlated with a higher destabilizing effect, while a positive value is associated with a variant predicted as stabilizing.

**Figure 1.**
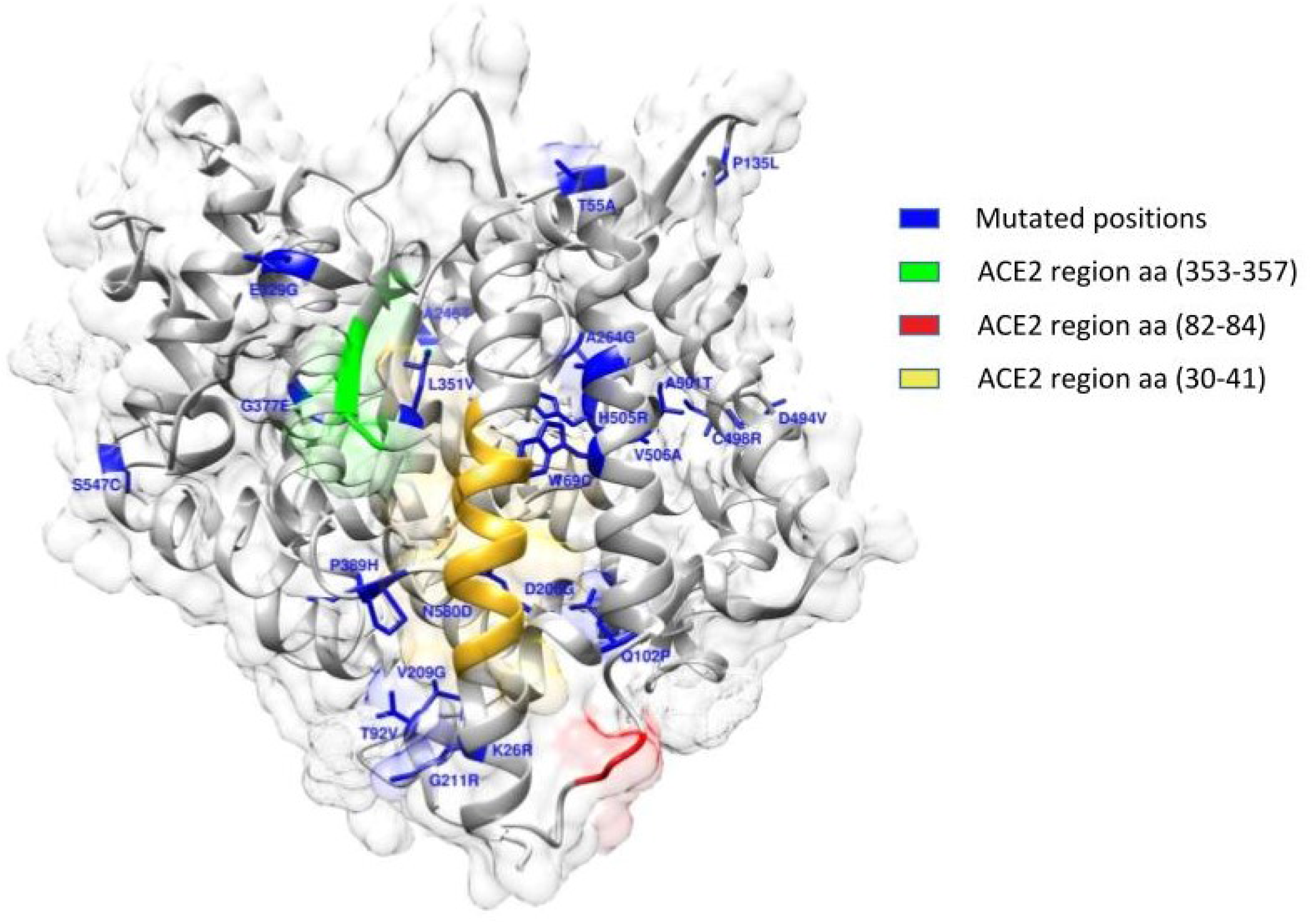
ACE2 Crystal structure with PDB ID 1R42. Surface and cartoon representations of protein in grey. In blue stick are represented each single mutated positions, cartoon region in yellow represent segment between amino acid 30-41, cartoon in green represent segment between amino acid 353-357 and cartoon in red represent segment between amino acid 82-84 that are protein regions responsible of interaction with 2019-nCOv spike glycoprotein

### ACE2 variants likely affect protein stability and SARS-CoV-2 binding

Molecular Dynamic analysis provides bona-fide simulations of protein structural changes caused by missense variants effects. Yet, its computationally expensive procedure led us to perform MD simulation for only a selection of representative candidate variants. Indeed, we selected the following five variants and corresponding effects: c.1517T>C p.(Val506Ala) which has the higher destabilizing effect, c.77A>G p.(Lys26Arg) and c.631G>A p.(Gly211Arg) with higher allele frequency along with c.1051C>G p.(Leu351Val) and the c.1166C>A p.(Pro389His) with a predicted effect on spike protein interaction. To analyse differences in protein structure between wild type and mutants, we performed 100ns MD simulations. The comparison was performed by Root Mean Square Deviation (RMSD) analysis. The global effects of the residue substitutions on flexibility and global correlated motion of ACE2 protein are represented in Figure 2 and the simulation is provided in Supplementary Video S1, S2, S3, S4, S5. While a similar trend for wild-type, c.77A>G p.(Lys26Arg) and c.1517T>C p.(Val506Ala) was observed with a steady course in the RMSD value, which stabilizes at an average of 0.2 nm, 0.25 nm and 0.3 nm respectively (Figure 3, panel A), the c.1166C>A p.(Pro389His) and c.1051C>G p.(Leu351Val) variants show a difference in comparison with the native protein with a gradual increase in RMSD value, which stabilizes at an average of 0.5 nm (Figure 3, panel A). Finally, the c.631G>A p.(Gly211Arg) shows a bigger difference with a higher increase in RMSD value, which stabilizes at an average of 0.6 nm (Figure 3, panel A). Structural analysis between WT and mutant c.1517T>C p.(Val506Ala) MD simulations showed that the c.1517T>C p.(Val506Ala) forms a hydrophobic centre together with Leu456, Leu503 and Phe516 with minimum differences in protein rearrangements when the residue is mutated in Ala as reported in Figure 2, Supplementary Video S1. The c.77A>G p.(Lys26Arg) is located at the N-terminus and the sidechain engages a hydrogen bond with Asn90 thus determining a minimal destabilizing predicted effect as shown in Table 2 and confirmed by RMSD analysis. The c.1166C>A p.(Pro389His) and the c.1051C>G p.(Leu351Val) variants, located in the region for the spike protein interaction, are characterized by a partially overlapping destabilizing effect. The c.1166C>A p.(Pro389His) variant sidechain being more bulky causes the shift of ACE2 (30-40) helix involved in spike protein interaction which being freer to move engages an interaction with Gln96 (Figure 2, Supplementary video S5). The c.1051C>G p.(Leu351Val) shorter sidechain is enable to interact with the hydrophobic core composed by Trp349 and Leu45 with a consequent rearrangement of the protein conformation. Finally, while c.631G>A p.(Gly211Arg) has theoretically a smaller destabilizing effect because of an external sidechain which is not involved in particular interaction network, as shown by MD simulation, it confers a wide flexibility to this region because the polar sidechain is able to engage different interactions with vicinal amino acid residues (Figure 2, Supplementary Video S2). During MD simulations, we have also investigated the surrounding region of ACE2 WT and previously selected variants by calculating change in Solvent Accessibility Surface Area (SASA). Differences in average SASA value would suggest for the native protein a wider surface exposed to solvent and subsequently a different ability to interact with spike SARS-CoV-2 in comparison with the studied variants (Figure 3, panel B).

**Figure 2.**
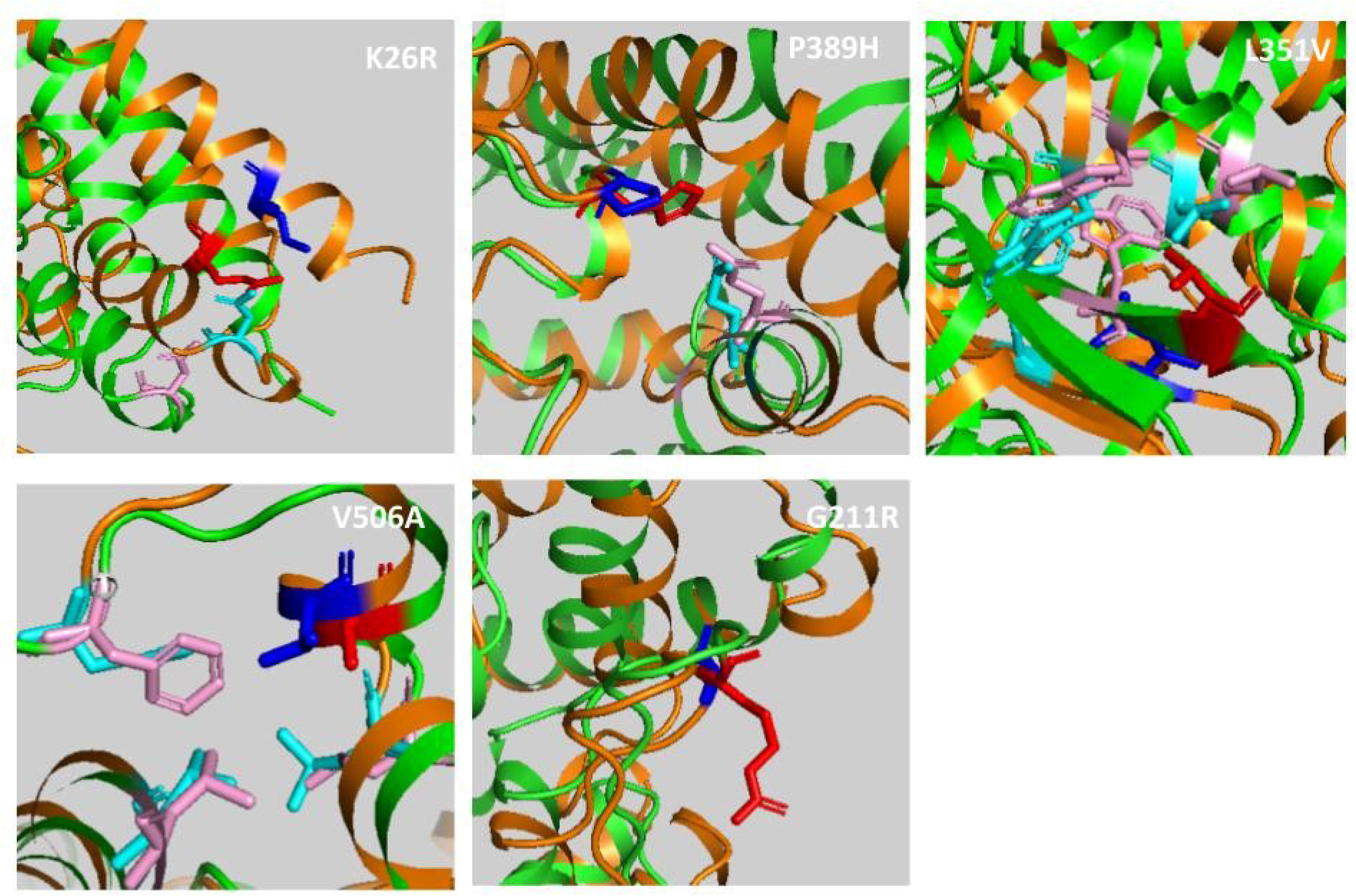
ACE2 wild type and variants superimposed structures after 100ns MD simulation. Cartoon representation of ACE2 wild type (orange) and variants (green) in blue sticks the wild type residues while in red the corresponding variants. In cyan and pink sticks residues interacting with each specific position.

**Figure 3:**
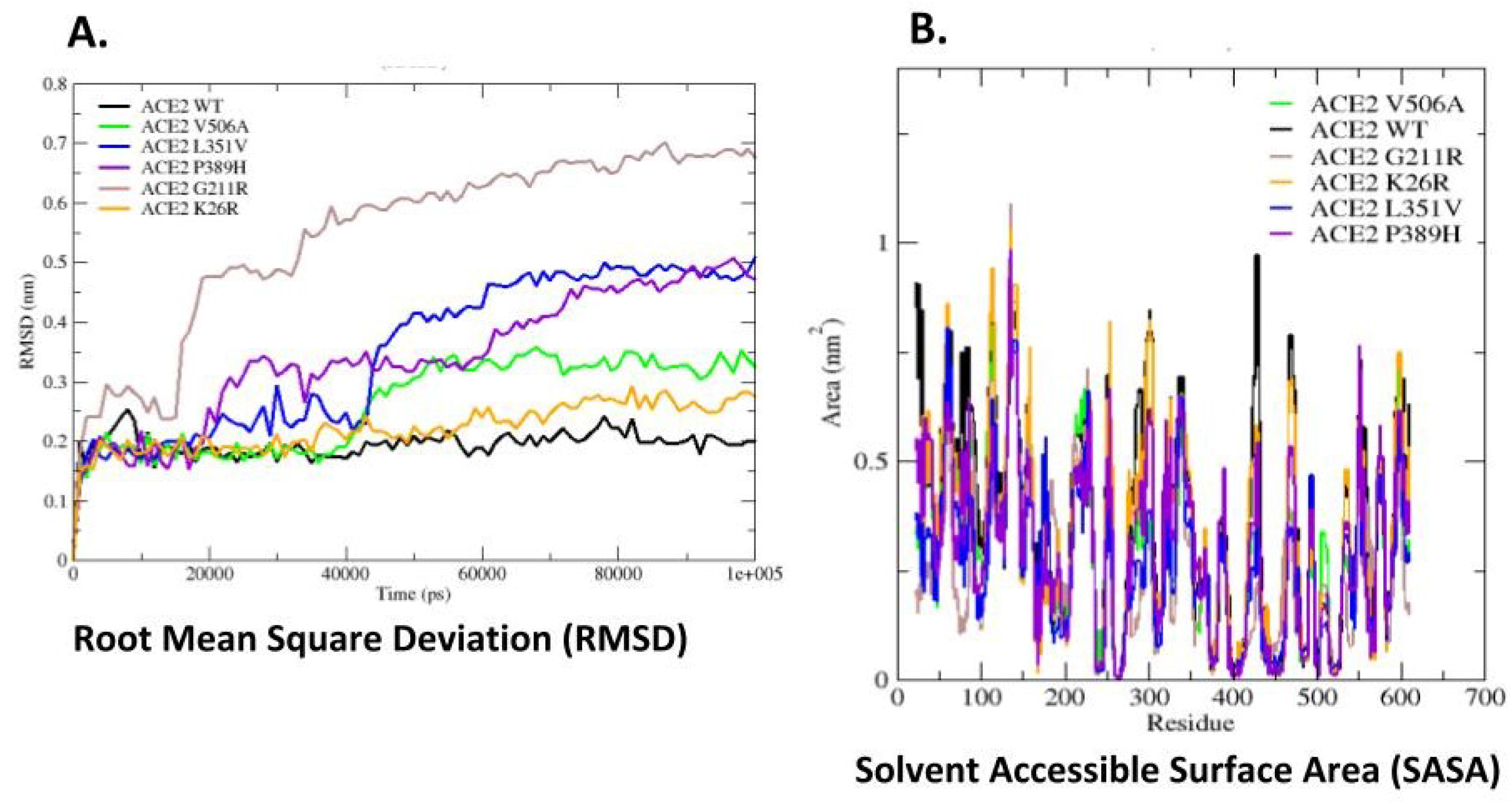
Structure superimposition snapshot between wild type protein and variant proteins. **Panel A**. Root Mean Square Deviation (RMSD) trends for the backbone of ACE2 WT (Black line) and some selected variants (colored lines, see legend) during 100 ns of simulation. The molecular dynamics simulation shows a good stability for all systems with exception of G211R mutants. (RMSD) is a parameter used to define the stability of an element. Wild type shows a steady course in the RMSD value, stabilizing at an average of 0.2 nm, while, the G211R variant shows a gradual increase in RMSD value, stabilizing at an average of 0.6 nm. **Panel B**. SASA graphical representation of ACE2 WT (Black line) and ACE2 variants (colored lines, see legend).

### Differences in *ACE2* allelic variability in COVID-19 patients compared to controls

In order to shed light on the role of ACE2 variants on interindividual variability and susceptibility to COVID-19 in Italian population we performed WES analysis on a cohort of 131 patients and 258 controls who agreed in participating to the study (see Materials and Methods). Data analysis of *ACE2* variants identified a different distribution of variants in controls compared to patients (Figure 4) with the c.2158A>G p.(Asn720Asp) variant being present in two hemizygous male patients (allele frequency 0,012) compared to 7 heterozygous female and 4 hemizygous male controls (allele frequency 0,028). A silent variant the c.2247G>A p.V749V, was also detected in 26 control individuals (allele frequency 0,069) compared to 5 COVID-19 patients (allele frequency 0,030). Although any single variant was not statistically significantly enriched in one cohort compared to the other, a cumulative analysis of the identified variants detected a statistically significant higher *ACE2* allelic variability (P value<0,029) in the control group compared to the patient cohort.

**Figure 4.**
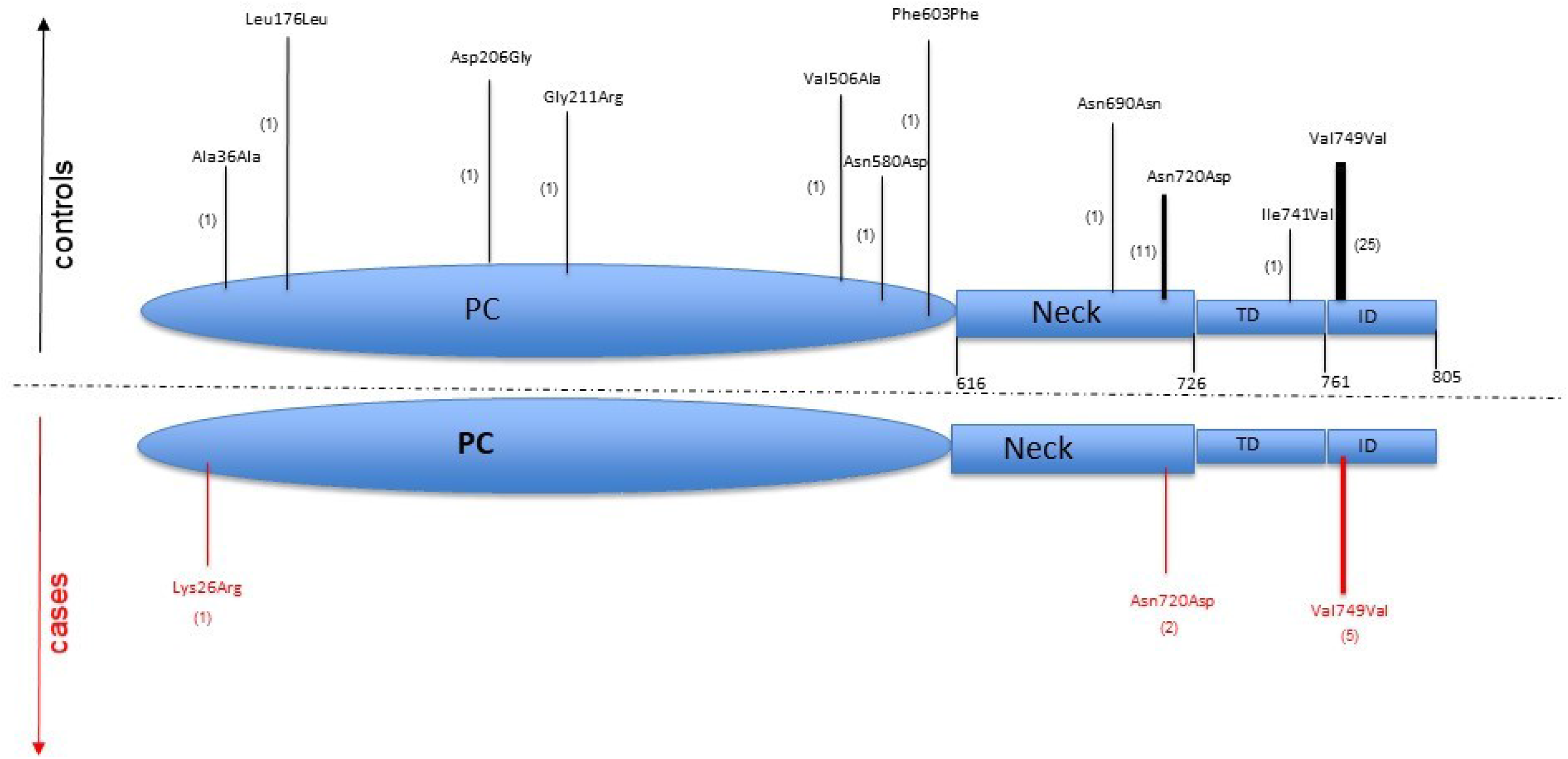
Differences in ACE2 variants in COVID-19 patients compared to controls. The figure shows the variants located in the ACE2 protein domains. The variants present in controls are shown in black while the variants in cases are shown in red. The number of patients carrying the variant is shown in brackets.

## DISCUSSION

According to recent reports, ACE2 is essential for SARS-CoV-2 to enter cells. Recent single-cell RNA studies have also shown that ACE2 is expressed in human lung cells (30). The majority of ACE2-expressing cells are alveolar type 2 (AT2) cells. Other ACE2 expressing cells include alveolar type 1 (AT1) cells, airway epithelial cells, fibroblasts, endothelial cells, and macrophages although their ACE2-expressing cell ratio is low and variable among individuals. The expression and distribution of the ACE2 receptor can thus justify the route of infection and the main localization at the alveolar level. Although the different density of ACE2 receptors in the upper respiratory tract among individuals can partially give reason for the clinical variability, which ranges from asymptomatic/paucisymptomatic patients to severely affected ones, it could not be the only reason for such variability. In addition, recent works did not observe significantly different viral loads in nasal swabs between symptomatic and asymptomatic patients (31). Italy has been the first European country that experienced the COVID-19 outbreak with a rapid increase in the positive cases in a very short-time period and a morbidity and lethality (~10%) definitely higher in comparison with Asian countries, such as China (4%) and South Korea (1.2%) (How deadly is COVID-19? A rigorous analysis of excess mortality and age-dependent fatality rates in Italy (32). These considerations raise the possibility of a predisposing genetic background accounting for or contributing to the wide inter-individual clinical variability, as well for the differential morbidity and lethality observed among different countries, population awareness and constrictive measures apart.

We integrated genomic WES data produced by five Italian centers (Siena, Naple, Turin, Bologna, Rome) interconnected by the Network for Italian Genome (NIG) in the attempt to identify variation encompassing the *ACE2* gene, which could account for a difference in SARS-CoV-2 spike binding affinity, processing or internalization. Previous studies showed that the residues near lysine 31, and tyrosine 41, 82-84, and 353-357 in human ACE2 are important for the binding of S-protein to coronavirus (12). In line with previous reports (33), we did not find polymorphism or rare variants in these residues in the Italian population. However, we identified three variants namely c.2158A>G p.(Asn720Asp), c.1166C>A p.(Pro389His) and c.1051C>G p.(Leu351Val), one of which polymorphic c.2158A>G (p.(Asn720Asp), moderately expressed in the Italian and European-non Finnish populations and with a very low allele frequency or not occurring in the Eastern Asia population. These variants which surround residual essentials for the SARS-CoV-2 spike protein binding were predicted to likely affect the cleavage dependent virion intake, such as the polymorphic c.2158A>G p.(Asn720Asp) (allele frequency 0.011) which lies 4 amino acids from the cleavage sequence of TMPRSS2 or to have a substantial impact on protein structure and spike protein interaction by MD simulation (Figure 3, Panel A). The relatively frequent c.631G>A p.(Gly211Arg) (allele frequency 0.0012, 12 /6930 individuals) was predicted to confer a wide flexibility to the region because of the ability to engage different interactions with the nearby amino acid residues. Along with these more common variants we also identified very rare variants such as the c.1166C>A p.(Pro389His) and the c.1051C>G p.(Leu351Val), some of which only described in the non-Finnish European population, that could give reason for a different affinity for the SARS-CoV-2 spike protein (Figure 2, Figure 3 panel A, Supplementary Video S4). Interestingly all the studied variants affect residues highly conserved among species (Supplementary Figure S1). Given their rarity in other populations, we cannot exclude that these variants can partially account for the clinical outcome observed in the Italian population. WES data generated from a wide cohort of COVID-19 Italian patients revealed a statistically significant (P<0,029) higher allelic heterogeneity for *ACE2* in controls compared to patients with a higher chance to find at least one *ACE2* variant in the cohort of controls compared to the cohort of patients. Therefore, it is plausible to think that the effect of allelic variability on ACE2 conformation would at least partially account for the interindividual clinical differences and likely modulate clinical severity. This finding reinforces the hypothesis that at least some of the identified variants or the cumulative effect of few of them confer a different susceptibility to virus cell entry and consequently to disease onset and progression. We cannot exclude that also silent variants such as the c.2247G>A (p.Val749Val) with no effect on the protein could play a role because of an unpredictable impact at a posttranscriptional level.

Notably, morbidity and lethality have been reported definitely higher in men compared to women (~70% vs 30%, 20th March 2020 ISS report). Although several parameters have been brought to case to explain this difference, i.e. smoking, differences in ACE2 localization and/or density in alveolar cells, hormonal asset, it is noteworthy that *ACE2* is located on chromosome X and that given the low allele frequency of the identified variants the rate of homozygous women is extremely low (see Results section). The X-chromosome inactivation (XCI) is incomplete in humans and some genes show a degree of XCI escape which vary between individuals and tissues (34). *ACE2* is one of the genes escaping X inactivation, but it belongs to a subgroup of X-chromosome genes showing a higher expression in men in several tissues thus mostly suggesting that ACE2 gene XCI is present although different in distinct tissues (15). Therefore, the impact of X-inactivation on the alternate expression of the two alleles would guarantee, in the affected tissues, a heterogeneous population of ACE2 molecules, some of which protective towards the infection until the point of a complete or almost complete protection in the case of a X-inactivation skewed towards the less SARS-CoV-2 -binding prone allele. This hypothesis would justify the high rate of asymptomatic or paucisymptomatic patients. However, the presented data does not allow to confirm a clear cause-effect relationship and, since most of the identified variants have very low frequencies, further functional studies are needed to validate these results. *ACE2* is definitely one of the main molecules whose genetic heterogeneity can modulate infection and disease progression; however, a deeper characterisation of the host genetics and functional variants in other pathway-related genes may help in understanding the pathophysiology of the disease opening up the way to a stratified risk assessment and to tailored preventive measures and treatments.

## Data Availability

I declare availability to the data referred to in the manuscript and note links below.

## Acknowledgements

This work was, in part, supported by: Telethon Network of Genetic Biobanks (project no. GTB12001), funded by Telethon Italy; Fondazione Bambino Gesu *(Vite Coraggiose* to M.T.); Mount Sinai, NY (USA) in the context of the international project ASC (Autism sequencing consortium); SOLVE-RD and MIUR project “Dipartimenti di Eccellenza 2018 – 2022” (n° D15D18000410001) to the Department of Medical Sciences, University of Torino (G.M., A.B.). We thank the CINECA consortium for providing computational resources.

## Ethical Approval

The NIG study was approved by the Ethical Committee of each center, Prot Name NIG Prot n 10547_2016, approved CEAVSE on 18.07.2016 n 88/16. The COVID-19 patients study was approved by the University Hospital (Azienda Ospedaliera Universitaria Senese) Ethical Committee, Siena, Italy (Prot n. 16929, dated March 16, 2020).

## Author’s contribution

EB, RT, OS, AMP and AR have made substantial contributions to conception and design, acquisition of data, analysis and interpretation of data and have been involved in drafting the manuscript. RA, GB, AB, AB, AT, GD, AG, FM, TP, AT, AT and FV has made substantial contributions to acquisition and analysis of the data. MB, MB, AC, SF, FM, GM, VN, MS and MT have made substantial contributions to interpretation of data and clinical evaluation. All authors have been involved in drafting the manuscript; have given final approval of the version to be published and agree to be accountable for all aspects of the work in ensuring that questions related to the accuracy or integrity of any part of the work are appropriately investigated and resolved.

---

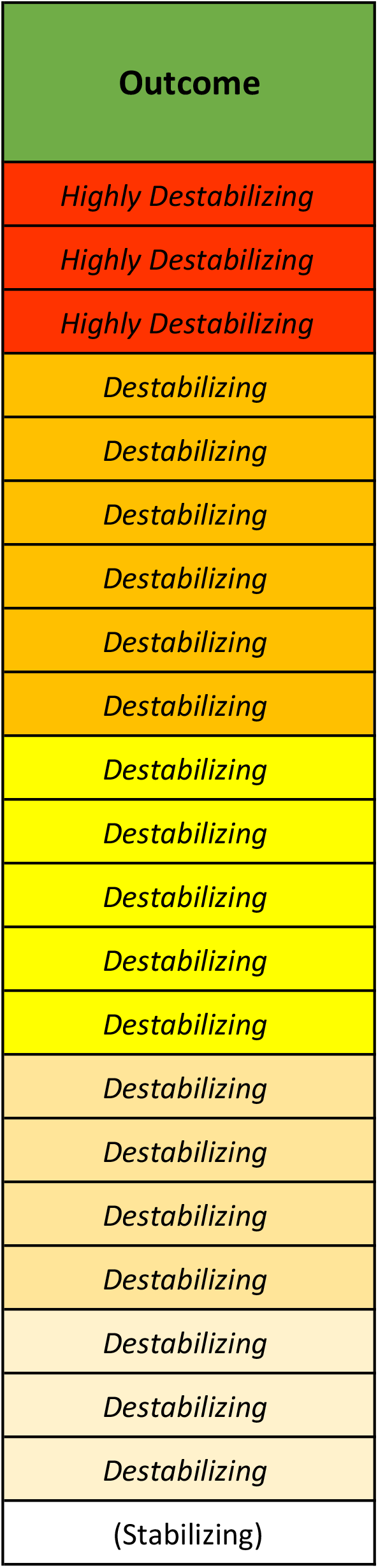

## References

1. Zhu N, Zhang D, Wang W, Li X, Yang B, Song J, et al. A novel coronavirus from patients with pneumonia in China, 2019. N Engl J Med. 2020;

2. Huang C, Wang Y, Li X, Ren L, Zhao J, Hu Y, et al. Clinical features of patients infected with 2019 novel coronavirus in Wuhan, China. Lancet. 2020;

3. Wang C, Horby PW, Hayden FG, Gao GF. A novel coronavirus outbreak of global health concern. The Lancet. 2020.

4. Chan JFW, Yuan S, Kok KH, To KKW, Chu H, Yang J, et al. A familial cluster of pneumonia associated with the 2019 novel coronavirus indicating person-to-person transmission: a study of a family cluster. Lancet. 2020;

5. Lai C-C, Liu YH, Wang C-Y, Wang Y-H, Hsueh S-C, Yen M-Y, et al. Asymptomatic carrier state, acute respiratory disease, and pneumonia due to severe acute respiratory syndrome coronavirus 2 (SARS-CoV-2): Facts and myths. J Microbiol Immunol Infect. 2020;

6. Zhou Y, Hou Y, Shen J, Huang Y, Martin W, Cheng F. Network-based drug repurposing for novel coronavirus 2019-nCoV/SARS-CoV-2. Cell Discov. 2020;

7. Xu Z, Shi L, Wang Y, Zhang J, Huang L, Zhang C, et al. Pathological findings of COVID-19 associated with acute respiratory distress syndrome. Lancet Respir Med. 2020;

8. Liu Y, Yan L-M, Wan L, Xiang T-X, Le A, Liu J-M, et al. Viral dynamics in mild and severe cases of COVID-19. Lancet Infect Dis [Internet]. 2020;2019(20):2019–20. Available from: https://doi.org/10.1016/S1473-3099(20)30232-2

9. Zhou P, Yang X-L, Wang X-G, Hu B, Zhang L, Zhang W, et al. A pneumonia outbreak associated with a new coronavirus of probable bat origin. Nature. 2020;

10. Wrapp D, Wang N, Corbett KS, Goldsmith JA, Hsieh C-L, Abiona O, et al. Cryo-EM structure of the 2019-nCoV spike in the prefusion conformation. Science (80-). 2020;

11. Yan R, Zhang Y, Li Y, Xia L, Guo Y, Zhou Q. Structural basis for the recognition of the SARS-CoV-2 by full-length human ACE2. Science. 2020;

12. Li W, Kuhn JH, Moore MJ, Wong SK, Huang IC, Farzan M, et al. Receptor and viral determinants of SARS-coronavirus adaptation to human ACE2. EMBO J. 2005;

13. Shulla A, Heald-Sargent T, Subramanya G, Zhao J, Perlman S, Gallagher T. A Transmembrane Serine Protease Is Linked to the Severe Acute Respiratory Syndrome Coronavirus Receptor and Activates Virus Entry. J Virol. 2011;

14. Heurich A, Hofmann-Winkler H, Gierer S, Liepold T, Jahn O, Pohlmann S. TMPRSS2 and ADAM17 Cleave ACE2 Differentially and Only Proteolysis by TMPRSS2 Augments Entry Driven by the Severe Acute Respiratory Syndrome Coronavirus Spike Protein. J Virol. 2014;

15. Tukiainen T, Villani AC, Yen A, Rivas MA, Marshall JL, Satija R, et al. Landscape of X chromosome inactivation across human tissues. Nature. 2017;

16. Benetti E, Giliberti A, Emiliozzi A, Valentino F, Bergantini L, Fallerini C, et al. Clinical and molecular characterization of COVID-19 hospitalized patients. medRxiv [Internet]. 2020 Jan 1;2020.05.22.20108845. Available from: http://medrxiv.org/content/early/2020/05/25/2020.05.22.20108845.abstract;

17. Magini P, Smits DJ, Vandervore L, Schot R, Columbaro M, Kasteleijn E, et al. Loss of SMPD4 Causes a Developmental Disorder Characterized by Microcephaly and Congenital Arthrogryposis. Am J Hum Genet. 2019;

18. Del Dotto V, Ullah F, Di Meo I, Magini P, Gusic M, Maresca A, et al. SSBP1 mutations cause mtDNA depletion underlying a complex optic atrophy disorder. J Clin Invest. 2020;

19. Flex E, Martinelli S, Van Dijck A, Ciolfi A, Cecchetti S, Coluzzi E, et al. Aberrant Function of the C-Terminal Tail of HIST1H1E Accelerates Cellular Senescence and Causes Premature Aging. Am J Hum Genet. 2019;

20. McKenna A, Hanna M, Banks E, Sivachenko A, Cibulskis K, Kernytsky A, et al. The genome analysis toolkit: A MapReduce framework for analyzing next-generation DNA sequencing data. Genome Res. 2010;

21. Wang K, Li M, Hakonarson H. ANNOVAR: Functional annotation of genetic variants from high-throughput sequencing data. Nucleic Acids Res. 2010;

22. McLaren W, Gil L, Hunt SE, Riat HS, Ritchie GRS, Thormann A, et al. The Ensembl Variant Effect Predictor. Genome Biol. 2016;

23. Rehm HL, Bale SJ, Bayrak-Toydemir P, Berg JS, Brown KK, Deignan JL, et al. ACMG clinical laboratory standards for next-generation sequencing. Genet Med. 2013;

24. Towler P, Staker B, Prasad SG, Menon S, Tang J, Parsons T, et al. ACE2 X-Ray Structures Reveal a Large Hinge-bending Motion Important for Inhibitor Binding and Catalysis. J Biol Chem. 2004;

25. Pires DEV, Ascher DB, Blundell TL. DUET: A server for predicting effects of mutations on protein stability using an integrated computational approach. Nucleic Acids Res. 2014;

26. Abraham MJ, Murtola T, Schulz R, Pall S, Smith JC, Hess B, et al. Gromacs: High performance molecular simulations through multi-level parallelism from laptops to supercomputers. SoftwareX. 2015;

27. Turner P. XMGRACE, Version 5.1. 19. Cent Coast Land-Margin Res Oregon Grad Inst Sci Technol Beavert. 2005;

28. Bussi G, Donadio D, Parrinello M. Canonical sampling through velocity rescaling. J Chem Phys. 2007;

29. Berendsen HJC, Postma JPM, Van Gunsteren WF, Dinola A, Haak JR. Molecular dynamics with coupling to an external bath. J Chem Phys. 1984;

30. Zhao Y, Zhao Z, Wang Y, Zhou Y, Ma Y, Zuo W. Single-cell RNA expression profiling of ACE2, the putative receptor of Wuhan 2019-nCov. bioRxiv. 2020;

31. D C, M T, F R, V D, M A, P P, et al. The early phase of the COVID-19 outbreak in Lombardy, Italy. 2020; Available from: http://arxiv.org/abs/2003.09320

32. Modi C, Boehm V, Ferraro S, Stein G, Seljak U. How deadly is COVID-19? A rigorous analysis of excess mortality and age-dependent fatality rates in Italy. medRxiv. 2020;

33. Cao Y, Li L, Feng Z, Wan S, Huang P, Sun X, et al. Comparative genetic analysis of the novel coronavirus (2019-nCoV/SARS-CoV-2) receptor ACE2 in different populations. Cell Discovery. 2020.

34. Carrel L, Willard HF. X-inactivation profile reveals extensive variability in X-linked gene expression in females. Nature. 2005;

